# Altered early cortical EEG maturation and its relationship to language development in Down syndrome

**DOI:** 10.64898/2026.08.16.26360527

**Authors:** Meagan Tsou, Haerin Chung, Katherine Pawlowski, Nicole Baumer, Carol Wilkinson

## Abstract

Down syndrome (DS) is the most common genetic cause of intellectual disability, yet age-related cortical maturation patterns that contribute to developmental delays remain poorly understood. We analyzed longitudinal resting-state EEG and developmental data from 86 children with DS and 154 typically developing (TD) children between 12 and 81 months of age. Linear mixed-effect models tested age-related trajectories of aperiodic and periodic spectral features, and manifold learning was used to characterize multivariate EEG profiles associated with age and developmental ability. Children with DS showed altered maturation across multiple EEG features. Aperiodic exponent decreased with age in DS, but not TD children, indicating possible altered maturation of cortical excitability. While TD children showed expected age-related increases in theta-alpha peak frequency and amplitude, children with DS exhibited limited alpha maturation and greater persistence of theta-only and theta+alpha peak profiles. We next asked whether multivariate EEG organization reflected chronological maturation or individual differences in developmental ability. A spectral dimension associated with chronological age in TD children was not similarly age-associated in DS. Instead, a second spectral dimension was associated with verbal developmental quotient in children with DS, independent of chronological age and nonverbal developmental ability. This language-associated profile included features considered atypical relative to TD maturation, including increased aperiodic activity and continued presence of theta activity. These findings suggest that in DS there is an altered relationship between cortical spectral organization, chronological age, and language development, extending beyond a uniform delay in typical maturation.

**Significance Statement:** Brain development in children with Down syndrome is often interpreted as delayed progression along a typical developmental path. Using longitudinal EEG across early childhood, we found that cortical spectral activity changed differently with age in Down syndrome. Moreover, spectral features that appeared less mature relative to typical development were associated with stronger language abilities within Down syndrome. These findings indicate that neural differences in Down syndrome do not solely reflect delay along a typical developmental timetable but may also represent distinct patterns of brain organization associated with developmental progress. Distinguishing altered organization from maturational delay is important for interpreting neural measures in Down syndrome and may help explain why developmental outcomes vary substantially among children with the same genetic condition.

## Introduction

Early brain development depends on the coordinated maturation of interacting neural systems. Synaptic development, inhibitory circuit maturation, thalamocortical signaling, and long-range network formation jointly shape the emergence of cortical rhythmic activity across infancy and childhood^1–5^. Genetic neurodevelopmental conditions may alter these coordinated processes at multiple points in development, resulting in neural trajectories that differ from typical development^6–8^. Some differences may reflect delayed or disrupted maturation, whereas others may reflect adaptation within an altered neural system^9^. Distinguishing among these possibilities is essential for understanding how genetic alterations influence brain development and contribute to variation in developmental outcomes.

Down syndrome (DS), caused by partial or complete triplication of chromosome 21, offers a genetically defined setting for examining how altered developmental cascades unfold in early development^10^. Although DS is the most common genetic cause of intellectual disability, developmental outcomes are highly variable across children^11^. Language abilities are particularly heterogenous and have major implications for learning, social communication, adaptive functioning, and long-term independence^12^. However, relatively little is understood about the early neurobiological pathways that contribute to developmental challenges and heterogeneity within DS. Improved understanding of alterations in early brain maturation in DS could help identify neural markers of developmental risk, clarify mechanisms underlying cognitive and language outcomes, and lay the groundwork for the development of new therapeutic interventions.

Resting-state electroencephalography (EEG) is a safe, non-invasive, and developmentally accessible method for studying early neural circuit maturation in infants and young children, including those with intellectual disability or limited task engagement^13^. The EEG power-spectrum summarizes ongoing neural activity across frequencies and can be separated into broadband aperiodic activity and frequency-specific periodic activity^14^. Aperiodic activity reflects the background organization of the spectrum and has been linked to features of cortical excitability and inhibition^15,16^, whereas periodic activity captures oscillations that arise from coordinated local and long-range circuit dynamics^4,17,18^. Together, these measures provide complementary information about cortical maturation and may help distinguish whether altered development in DS reflects a delay in expected maturational patterns, divergence from typical trajectories, or distinct neural organization associated with variation in developmental outcomes.

In typical development, EEG spectral features show organized age-related changes across infancy and early childhood^19–21^. Aperiodic activity increases rapidly during the first year of life, followed by relative stabilization and later age-related reduction across later childhood and adulthood^19,22–24^. Periodic features also exhibit dynamic developmental changes; theta/alpha peak profiles that may be present in early infancy become less common with age^19^, alpha rhythms increase in amplitude and peak frequency^25–27^, and low beta activity emerges across infancy^19,28^. Together, these patterns provide a developmental map to determine whether DS maturation follows expected age-related trajectories or diverges from them.

Prior EEG studies with older children and adults with DS have identified differences across several of these same spectral domains, most consistently involving increased slow frequency activity and reduced alpha activity^29–31^. However, relatively few studies have examined EEG spectral development in young children with DS. In our prior cross-sectional work, toddlers and preschoolers with DS showed reduced aperiodic slope, increased periodic theta power, reduced alpha peak amplitude, and reduced beta activity compared with age-matched and developmental-level-matched comparison groups^29^. Increased theta power was driven in part by the persistence of theta/alpha peak profiles that are typically observed only much earlier in development^19,29^. These findings suggest that EEG maturation in DS may include early-like spectral features, but longitudinal data are needed to determine whether these features change with age and whether multivariate patterns of spectral activity relate to variation in developmental outcomes.

In the present study, we asked whether EEG differences in Down syndrome reflect slower progression along typical maturational trajectories or a distinct organization of cortical spectral development. We address this question using longitudinal resting-state EEG data from a substantially larger cohort of children with DS than our prior cross-sectional study. We first examined whether age-related trajectories of periodic and aperiodic EEG features differed between children with DS and typically developing children. We then tested whether multivariate patterns of spectral activity related to chronological age or to individual differences in verbal and nonverbal developmental ability. We found that children with DS showed altered age-related spectral organization and that the relationships between multivariate EEG patterns, age, and developmental ability differed across groups. In typical development, one multivariate spectral dimension was strongly related to chronological age, whereas in Down syndrome a distinct spectral dimension was associated with verbal ability. Together, these findings support a model in which cortical spectral development in Down syndrome reflects altered developmental organization rather than a uniform delay in typical maturation.

## Results

The final analytical sample included 240 unique participants (DS: n = 86, TD: n = 154) and 448 resting-state EEG recordings (DS: n = 120, TD: 328). EEG recordings ranged from 12 to 81 months of age. Participant age distribution by group is shown in Figure 1, and demographic and developmental characteristics are summarized in Table 1. Groups differed in race, household income, and parent education, with lower household income and parent education in the DS group. Groups also differed on several EEG quality metrics, with higher amounts of artifact in EEGs collected from participants with DS (SI Appendix, Section S1.4).

**Figure 1.**
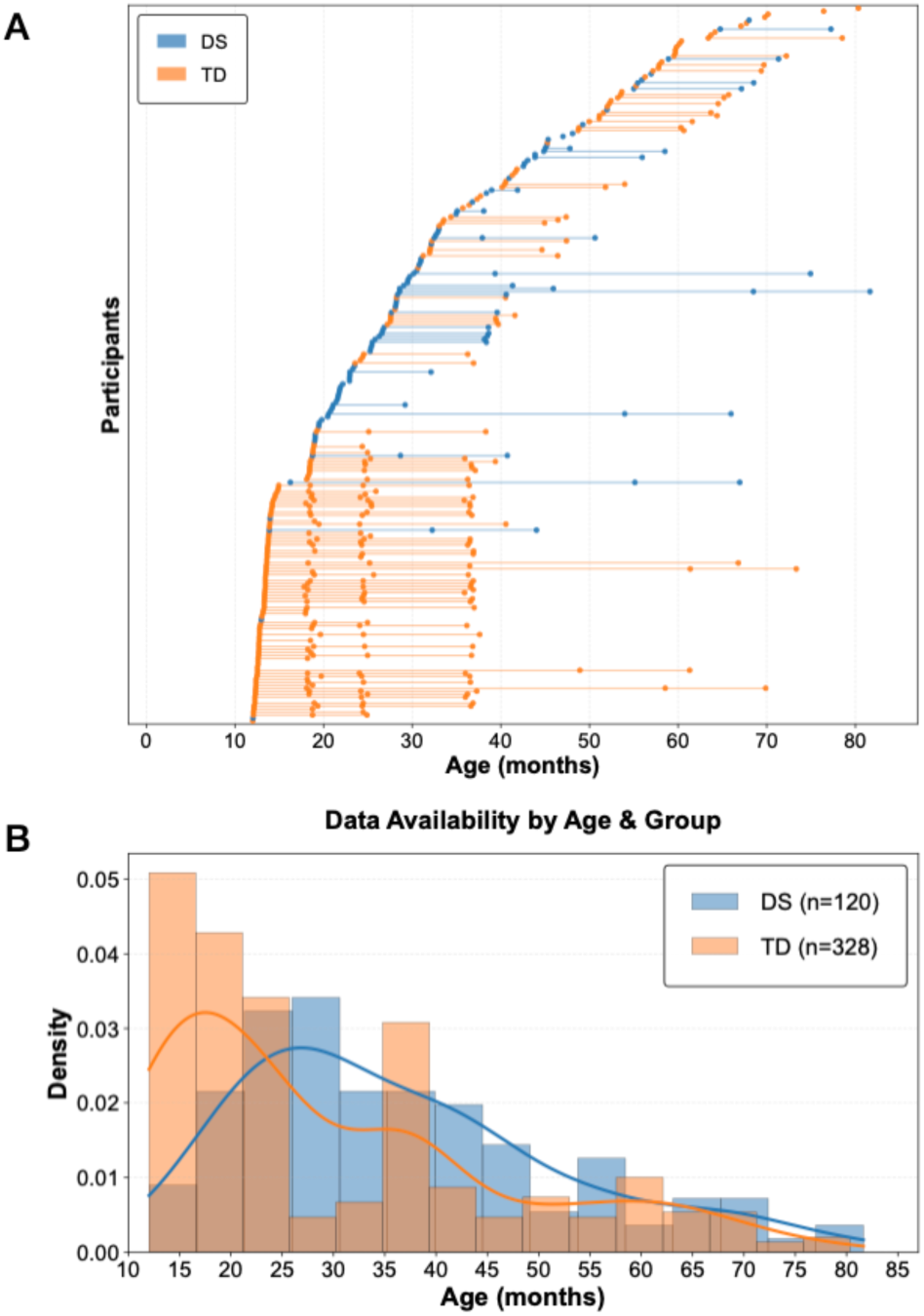
(A) Timeline showing EEG visits for individual Down Syndrome (DS; blue) and typically developing (TD; orange) participants across age. (B) Histogram and kernel density estimation showing the distribution of visits by age and group.

**Table 1:** Sample Characteristics.

| | DS (n=84)<br>n (%) | TD (n=154)<br>n (%) | $\chi^2(df)$ | p |
| --- | --- | --- | --- | --- |
| **Sex** | | | $\chi^2(1) = 0.169$ | 0.681 |
| Female | 40 (47) | 66 (43) |  |  |
| **Race** | | | $\chi^2(4) = 9.593$ | 0.048 |
| White | 73 (85) | 110 (71) |  |  |
| Black or African American | 4 (5) | 7 (5) |  |  |
| Asian | 1 (1) | 8 (5) |  |  |
| More than 1 race | 5 (6) | 27 (18) |  |  |
| Race was not listed | 1 (1) | 1 (1) |  |  |
| Missing/Prefer not to say | 2 (2) | 1 (1) |  |  |
| **Ethnicity** | | | $\chi^2(1) = 1.321$ | 0.25 |
| Hispanic or Latino | 10 (12) | 10 (6) |  |  |
| Missing / prefer not to say | 1 (1) | 1 (1) |  |  |
| **Household Income\*** | | | $\chi^2(2) = 17.45$ | <.001 |
| \$70,000 and greater | 66 (77) | 144 (94) | | |
| \$40,000 - \$69,999 | 12 (14) | 2 (1) | | |
| Less than \$40,000 | 2 (2) | 6 (4) | | |
| Missing or prefer not to disclose | 6 (7) | 2 (1) |  |  |
| **Highest Parent Education, n (%)** | | | $\chi^2(3) = 8.434$ | 0.038 |
| > 4-year college degree | 49 (57) | 114 (74) |  |  |
| Bachelor's degree | 25 (29) | 27 (18) |  |  |
| Associate degree | 6 (7) | 5 (3) |  |  |
| ≤ High school/GED | 6 (7) | 6 (4) |  |  |
| Missing or prefer not to disclose | - | 2 (1) |  |  |
| **Developmental Quotients, Mean (SD), [Range], N** |  |  |  |  |
| Verbal Developmental Quotient | 52.2 (13.7)<br>[23.96 - 89.47]<br>N = 85 | 109.7 (14.9)<br>[12.0-78.5]<br>N=309 |  |  |
| Nonverbal Developmental Quotient | 51.95 (13.7)<br>[8.89 - 81.25]<br>N = 85 | 112.7 (18.3)<br>[61.5 - 154.2]<br>N = 320 |  |  |

\*Income responses spanning multiple categories were assigned to the higher tier: "\$50,000–\$100,000" was assigned to "\$70,000 and greater" (n = 9), "Less than \$50,000" was assigned to "\$40,000–\$69,999" (n = 2), and "\$35,000–\$75,000" was assigned to "\$40,000–\$69,999" (n = 2) to maintain consistency with the broader harmonized values.

### Children with DS show altered age-related trajectories of EEG spectral features

EEG Spectral features showed distinct age-related patterns in children with DS relative to TD children. Absolute, aperiodic, and periodic spectra by diagnostic group and age bin are shown in Figure 2. Linear mixed-effects models were used to statistically assess age-related trajectories of EEG features (Table 2, Figure 3), with an EEG quality metric included as a covariate to account for group differences in quality. Models revealed no group differences in aperiodic offset at 36 months, but a marginally significant group × age interaction (q = 0.061), with a significant age-related decline in exponent in children with DS (p < .001) but not TD children (p = .073)(Figure 3B). This pattern suggests age-related flattening of the aperiodic spectrum in DS across the toddler and preschool age range.

**Figure 2.**
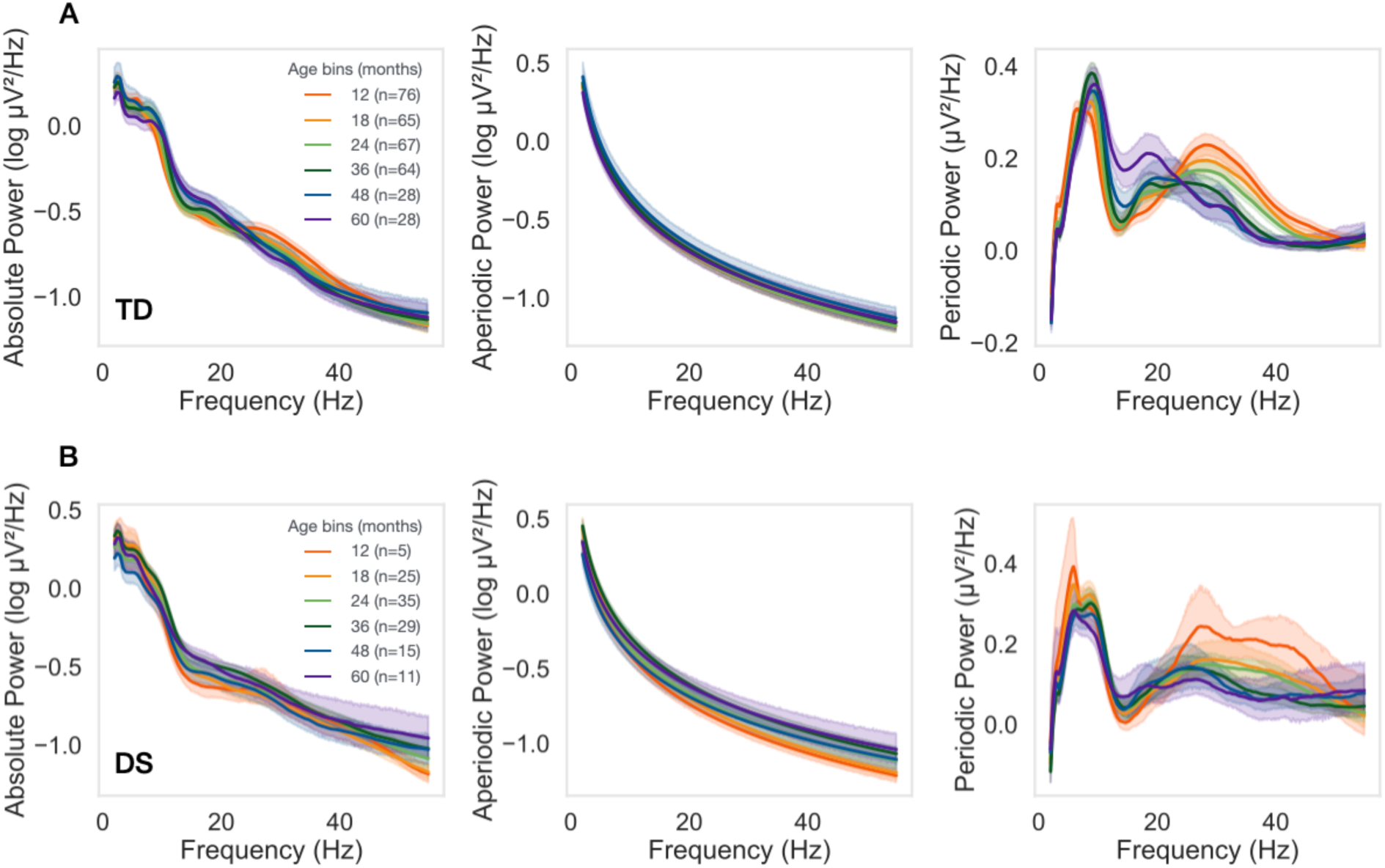
Power spectral density plots by age bins in children with TD (top) and DS children (bottom). Each panel displays mean absolute power (left), aperiodic power (middle) and periodic power (right) across frequencies. Mean values for each age bin are shown as colored lines with 95% confidence intervals (shaded regions).

**Figure 3.**
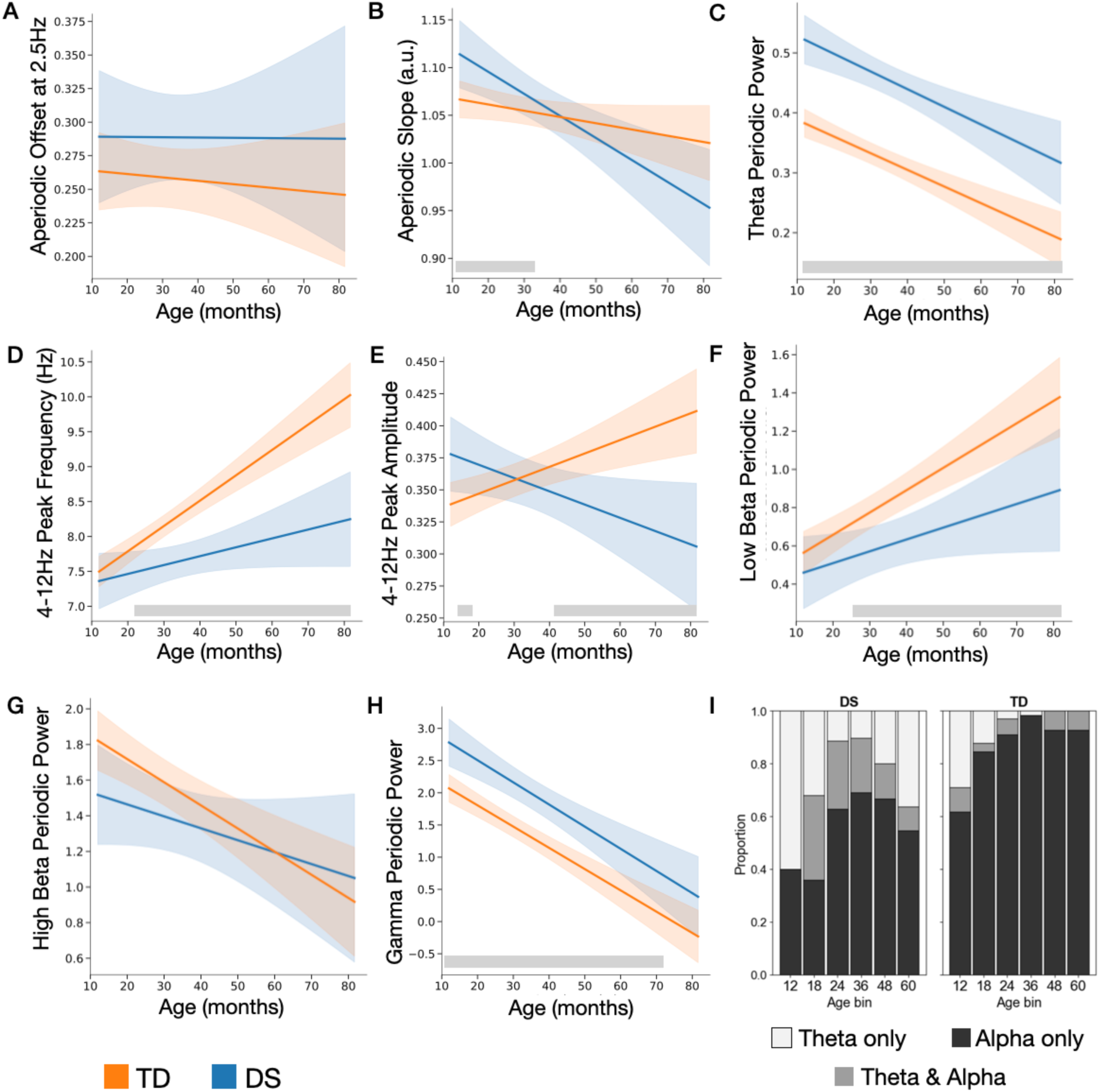
**(A-H).** Change in EEG measures across age in children with OS (blue) and typically developing children (orange). Lines are the model predicted value with the shaded area representing 95% confidence intervals. Gray shaded horizontal bars show age bands with significant difference between groups. (1) Proportion of children with either only theta, only alpha, or both theta and alpha peaks present between 4-12 Hz.

**Table 2:** Linear mixed effect models.

| | Group effect at 36m<br>$\beta$ [95% CI], $q$ | Group × Age<br>Interaction<br>$\beta$ [95% CI], $q$ | TD slope<br>$\beta$/year [95% CI], $p$ | DS slope<br>$\beta$/year [95% CI], $p$ |
| --- | --- | --- | --- | --- |
| EEG Feature |  |  |  |  |
| Aperiodic Offset | 0.03 [-0.01, 0.07]<br>$q = .190$ | 0.00 [-0.02, 0.03]<br>$q = .852$ | -0.00 [-0.01, 0.01]<br>$p = .609$ | -0.00 [-0.02, 0.02]<br>$p = .979$ |
| Aperiodic Exponent | 0.01 [-0.02, 0.03]<br>$q = .563$ | -0.02 [-0.04, -0.00]<br>$q = .061$ | -0.01 [-0.02, 0.00]<br>$p = .073$ | -0.03 [-0.04, -0.01]<br>**$p < .001$** |
| 4-12Hz Peak Frequency | -0.70 [-0.99, -0.41]<br>**$q < .0001$** | -0.28 [-0.48, -0.09]<br>**$q = .017$** | 0.44 [0.33, 0.54]<br>**$p < .0001$** | 0.15 [-0.01, 0.32]<br>$p = .070$ |
| 4-12Hz Peak Amplitude | -0.01 [-0.03, 0.01]<br>$q = .408$ | -0.02 [-0.04, -0.01]<br>**$q = .003$** | 0.01 [0.01, 0.02]<br>**$p < .001$** | -0.01 [-0.02, -0.00]<br>**$p = .038$** |
| Periodic Theta Power | 0.14 [0.10, 0.17]<br>**$q < .0001$** | -0.00 [-0.02, 0.02]<br>$q = .852$ | -0.03 [-0.04, -0.02]<br>**$p < .0001$** | -0.04 [-0.05, -0.02]<br>**$p < .0001$** |
| Periodic Low Beta Power | -0.24 [-0.39, -0.08]<br>**$q = .005$** | -0.07 [-0.15, 0.02]<br>$q = .291$ | 0.14 [0.10, 0.19]<br>**$p < .0001$** | 0.07 [-0.00, 0.15]<br>$p = .054$ |
| Periodic High Beta Power | -0.15 [-0.38, 0.07]<br>$q = .250$ | 0.08 [-0.05, 0.21]<br>$q = .409$ | -0.16 [-0.22, -0.09]<br>**$p < .0001$** | -0.08 [-0.19, 0.03]<br>$p = .156$ |
| Periodic Gamma Power | 0.68 [0.39, 0.97]<br>**$q < .0001$** | -0.02 [-0.19, 0.16]<br>$q = .852$ | -0.40 [-0.49, -0.31]<br>**$p < .0001$** | -0.41 [-0.56, -0.26]<br>**$p < .0001$** |

Periodic differences were also observed. Children with DS showed elevated periodic theta power relative to TD children across the age range (Fig 3C, q < .0001), and theta power decreased with age across both groups, with no significant group × age interaction. Peak-profile analyses further indicated that theta-dominant spectral profiles were more common in DS (Figure 3I). Between 24 and 48 months, when age distributions were more comparable across groups, theta-only or theta-plus-alpha profiles were observed in 32% of children with DS compared with 5% of TD children (χ²s ≥ 39.6, p = 2 × 10^-9). In addition, there were significant group differences in developmental trajectories of peak frequency and amplitude between 4 to 12hz. TD children showed significant age-related increases in peak frequency and amplitude, consistent with expected developmental increases in alpha organization. In contrast, children with DS showed no significant age-related association for peak frequency and a negative association for peak amplitude (Figure 3D,E). Thus, the expected age-related strengthening and acceleration of theta-alpha activity observed in TD children was not observed in children with DS.

Children with DS also exhibited differences in higher frequency bands. Specifically, they had reduced periodic low beta power relative to TD children after 23 months of age. Although the group × age interaction was not significant, low beta power increased significantly with age in TD children but not in children with DS. No significant group difference or group × age interaction was observed for periodic high beta power, although high beta power decreased with age in TD children and showed no significant age association in DS. Finally, across the age range, children with DS had increased periodic gamma power compared to TD counterparts (q<.0001).

Sensitivity analyses confirmed that including household income as a covariate in models did not change any of the above results (SI Appendix, Section S2.1). Analyses were also completed for across four electrode based regions of interests, with general consistency in trends across regions (SI Appendix, Section S2.2)

### EEG spectral dimensions show distinct associations with chronological age and language ability across groups

To examine the association between multivariate EEG activity and developmental outcomes in DS children we applied a three-step analysis. First, to reduce dimensionality and minimize collinearity among neighboring frequency bins, spectral power features were summarized into nine EEG-derived measures: the aperiodic exponent, periodic total power, periodic gamma power, and a categorical theta/alpha peak index, as well as the weights of five principal components derived the periodic spectrum between either 4-12hz (three components) or 12-30hz (two components) (SI Appendix, Section S1.6). Second, because individual EEG features may not adequately capture the altered developmental organization observed in DS, we used PHATE (Potential of Heat-diffusion for Affinity-based Transition Embedding^32^), an unsupervised manifold-learning method, to identify low-dimensional structure across multiple EEG features. Two PHATE dimensions were identified with feature weights shown in Figure 4A, and average spectra for individuals with low, middle, or high PHATE dimension scores are shown in Figure 4B. Finally, we then examined associations between PHATE-derived manifold scores, chronological age, and developmental ability. Verbal developmental quotient (VDQ), derived from receptive and expressive language measures from either the Mullen Scales of Early Learning (MSEL^33^) or Preschool Language Scale (PLS-5^34^) indexed language ability relative to chronological age. Nonverbal developmental quotient (NVDQ) was derived from the MSEL visual reception and fine motor subscales (SI Appendix, Section S1.5)

**Figure 4.**
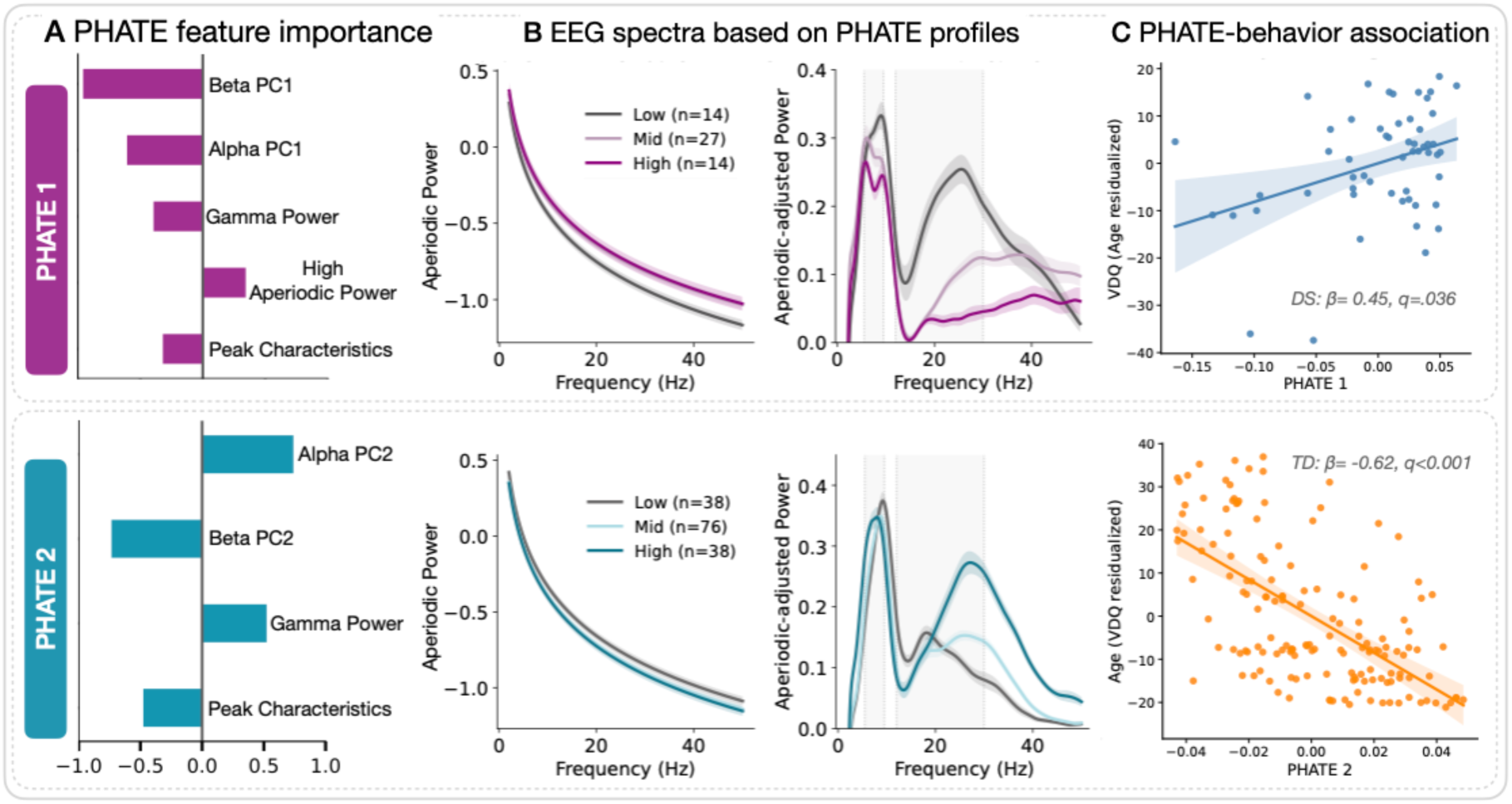

The two PHATE dimensions showed distinct group-specific associations with language development and chronological age (Table 3). PHATE 2 tracked chronological age, reflecting a developmental maturation axis in TD (β = -0.62, permutation q < .001) but not in DS (q = 0.226), with a significant PHATE 2 × Group interaction confirming that the age-related association differed between groups (β = 0.56, permutation p < .001). TD children with low PHATE 2 scores were older, and had lower aperiodic power, higher alpha peak frequency, and low beta power spectral profiles (Fig 4B,C). In contrast, PHATE 1 was associated with verbal skills only within the DS group accounting for age (β = 0.45, permutation q = 0.036), whereas no association was observed in the TD group (permutation q = .886). Consistent with this pattern, the PHATE1 × Group interaction was significant (β = 0.20, permutation p = .042), indicating that the relationship between PHATE1 and verbal ability differed between diagnostic groups. Children with DS with high PHATE 1 scores, had higher VDQ, high aperiodic activity, double theta-alpha peaks, and reduced beta spectral profiles (Fig 4B,C).

**Table 3:** Regression models of PHATE dimensions, chronological age, verbal ability (VDQ), and non-verbal ability (NVDQ).

| Predictor | $\beta$ (SE) | t | 95% CI | p | p (perm) | q (FDR) |
| --- | --- | --- | --- | --- | --- | --- |
| DS: PHATE1 ~ VDQ + Age (n = 55, $R^2 = 0.13$) | | | | | | |
| **VDQ** | **0.45 (0.16)** | **2.76** | **[0.12, 0.77]** | **.008** | **.009** | **.036** |
| Age | 0.22 (0.16) | 1.37 | [-0.10, 0.55] | .175 | .169 | .225 |
| TD: PHATE1 ~ VDQ + Age (n = 152, $R^2 = 0.02$) | | | | | | |
| VDQ | -0.01 (0.08) | -0.14 | [-0.17, 0.15] | .888 | .886 | .886 |
| Age | -0.14 (0.08) | -1.69 | [-0.30, 0.02] | .092 | .088 | .177 |
| DS: PHATE2 ~ VDQ + Age (n = 55, $R^2 = 0.03$) | | | | | | |
| VDQ | -0.04 (0.17) | -0.25 | [-0.38, 0.30] | .805 | .813 | .886 |
| Age | -0.21 (0.17) | -1.22 | [-0.55, 0.13] | .229 | .226 | .226 |
| TD: PHATE2 ~ VDQ + Age (n = 152, $R^2 = 0.38$) | | | | | | |
| VDQ | -0.04 (0.06) | -0.59 | [-0.17, 0.09] | .556 | .568 | .886 |
| **Age** | **-0.62 (0.06)** | **-9.65** | **[-0.75, -0.49]** | **<.001** | **<.001** | **<.001** |
| DS verbal specificity: VDQ ~ PHATE1 + NVDQ + Age (n = 55, $R^2 = 0.65$) | | | | | | |
| **PHATE 1** | **0.19 (0.08)** | **2.22** | **[0.02, 0.36]** | **.031** | **.030** |  |
| NVDQ | 0.59 (0.11) | 5.61 | [0.38, 0.80] | <.001 | <.001 |  |
| Age | -0.24 (0.10) | -2.27 | [-0.44, -0.03] | .027 | .031 |  |
| DS Non-verbal: NVDQ ~ PHATE1 + VDQ + Age (n = 55, $R^2 = 0.62$) | | | | | | |
| PHATE 1 | -0.02 (0.09) | -0.19 | [-0.20, 0.17] | .851 | .845 |  |
| VDQ | 0.65 (0.12) | 5.61 | [0.41, 0.88] | <.001 | <.001 |  |
| Age | -0.22 (0.11) | -1.97 | [-0.44, 0.00] | .054 | .050 |  |

To assess specificity, we tested whether PHATE 1 remained as a significant predictor when adjusting for chronological age and nonverbal developmental quotient. PHATE1 remained an independent predictor of verbal ability (β = 0.19, p = .031). In contrast, PHATE1 was not associated with nonverbal developmental quotient after adjusting for verbal ability (β = -0.02, p = .85), suggesting that the association was specific to verbal rather than general cognitive ability. This association was robust across different PHATE hyperparameters, suggesting that our findings reflect genuine low-dimensional spectral structure rather than a property of the PHATE algorithm (SI Appendix, Section S2.3).

## Discussion

In this study, we examined developmental trajectories of resting-state EEG spectral features in infants and young children with Down syndrome or typical development. Across feature-level and multivariate analyses, children with DS showed evidence of altered cortical maturation rather than uniform delay in typical developmental patterns. At the feature level, children with DS showed age-related flattening of the aperiodic spectrum, elevated periodic theta power, reduced maturation of 4-12 Hz peak frequency and amplitude, and a greater prevalence of theta-only and theta+alpha peak profiles. At the multivariate level, PHATE identified a spectral dimension associated with chronological age, but this age-related association was not present in DS. Instead, in DS, a different spectral dimension was associated with verbal developmental quotient, independent of both nonverbal skills and chronological age. Together, these findings suggest that trisomy 21 alters the developmental organization of cortical rhythmic activity, and that individual differences in spectral activity may relate to language development in DS more than to chronological age alone.

### Decreases in aperiodic exponent across early childhood in DS

We observed that aperiodic exponent declined with age in children with DS, suggesting age-related flattening of the power spectrum. A flatter aperiodic spectrum has been shown to reflect greater relative excitation or reduced inhibition^15,35,36^. This finding is notable given longstanding hypotheses regarding altered excitatory-inhibitory balance in DS. Much of the preclinical literature has emphasized GABAergic over-inhibition, supported by evidence of altered interneuron development, increased GABAergic signaling, and impaired synaptic plasticity circuits^37–39^. However, GABAergic signaling is developmentally complicated. Early in development, GABA first exerts depolarizing or excitatory effects before the maturation of chloride gradients that support hyperpolarizing inhibitory GABA signaling^40,41^. Studies in DS mouse models suggest that this excitatory-to-inhibitory transition may be altered or delayed^42,43^, raising the possibility that increased GABAergic signaling does not necessarily translate into greater functional inhibition at all developmental stages. In this context, the age-related flattening observed here may reflect altered maturation of cortical population activity in DS, potentially related to changes inhibitory signaling, or shifts in surface cortical excitability.

### Alteration in theta-alpha peak profiles in Down Syndrome

The altered theta and alpha peak profiles observed in DS may reflect disruption of a broader developmental transition in the organization of spontaneous EEG rhythms. In typical development, early lower-frequency rhythms shift to stable alpha range activity, reflected in age-related increases in both alpha peak frequency and amplitude^20,25,27^. This transition was evident in the TD group, but not in children with DS who showed persistent theta activity and a higher prevalence of theta-only and theta+alpha peak profiles. This pattern suggests an altered or incomplete theta-to-alpha transition, rather than a simple delay in alpha maturation. In typical early development, changes in dominant EEG rhythms may reflect maturation of thalamocortical circuitry, including the transition from early subplate-mediated thalamic input toward more mature cortical-cortical and corticothalamic loops^18,26^. Under this framework, persistent theta rhythms or simultaneous theta and alpha peaks in DS could indicate incomplete stabilization of mature alpha generating circuitry.

Several additional mechanisms could contribute to this altered theta-to-alpha transition. GABAergic maturation is likely relevant as inhibitory interneurons help regulate the timing and stability of cortical oscillations. Altered GABA polarity or chloride homeostasis could therefore contribute to both aperiodic flattening and persistence of immature theta-alpha organization. Recent single-cell multiomic work in developing Ts21 human neocortex also suggests altered timing and patterning of cortical neurogenesis, with fewer deep-layer corticothalamic neuron populations and relatively greater development of excitatory upper-layer intratelencephalic neurons^44^. Because mature alpha rhythms depend in part on reciprocal thalamocortical and corticothalamic feedback loops, disruption of the cortical projection neurons that support these loops could provide one mechanism for attenuated alpha maturation and persistent theta-alpha disorganization in DS. In this interpretation, altered inhibitory maturation may affect local cortical excitability and oscillatory timing, while altered corticothalamic development may impair the feedback pathways needed to stabilize alpha rhythms.

### EEG profiles and developmental outcomes

Our multivariate analyses further suggest that EEG spectral profiles have different developmental significance in children with DS and typical development. In TD children, PHATE identified a spectral dimension strong associated with chronological age, consistent with expected maturation of alpha and beta activity. In contrast, this age-related spectral profile association was disrupted in DS. Instead, a separate PHATE dimension was associated with verbal development quotient in children, even accounting for chronological and nonverbal developmental ability. This dissociation suggests that EEG variation in DS is not a weaker version of typical age-related maturation, but rather, spectral features that appear atypical relative to TD development may carry DS-specific developmental information.

The direction of DS language association was unexpected and may be informative. Higher verbal developmental quotient was associated with a multivariate profile that included increased aperiodic power, theta-alpha peaks, and reduced beta power. One possibility is that this profile reflects an alternative compensatory network organization that supports language development in the context of altered DS brain maturation. This interpretation is consistent with evidence from other neurodevelopmental conditions suggesting that atypical neural features may reflect compensatory or homeostatic adaptations. For example, in Fragile X Syndrome (FXS) increased aperiodic gamma power has been associated with better language skills^45^, with increased aperiodic gamma power potentially reflecting adaptive changes in network organization that help maintain firing rates^46^. In DS, the PHATE1 profile may similarly reflect variation in how cortical networks maintain function despite altered maturation of alpha generating circuitry.

A second, non-mutually exclusive explanation is that the EEG features associated with verbal ability in DS reflect, developmental level rather than chronological age. Some features contributing to PHATE1 and VDQ associations, including reduced beta power and altered theta-alpha organization, resemble patterns observed in earlier stages of typical development (approximately 2 to 18 months;^19^). This raises the possibility that EEG maturation in DS follows aspects of typical spectral sequence, but on a timescale more closely related to language ability than age. If this interpretation is correct, increased alpha power in DS does not necessarily indicate a more mature neural state. Alpha power reflects multiple interacting physiological properties including oscillatory amplitude, peak frequency, burst dynamics, and modulation of aperiodic dynamics^47^. Similar average alpha power measures can therefore arise from different underlying circuit dynamics. Future analyses of burst amplitude, duration, and temporal consistency, may help determine whether language-associated EEG profiles in DS reflect adaptive organization, delayed maturation indexed by developmental level, or distinct oscillatory dynamics.

### Limitations

This study has several limitations. While our sample size is large relative to prior EEG studies in young children with DS, the number of children with DS in the youngest age bin was limited, reducing our ability to assess differences at the very earliest times in development. In addition, our TD group had less sociodemographic diversity than our DS group. While findings remained the same when including household income as a covariate in statistical models, future studies need to improve recruitment of children from diverse backgrounds. Finally, given the age of the participants in the study, there is likely variability in the attentional state of each child that may influence resting state EEG activity.

## Conclusion

In summary, young children with DS showed altered developmental organization of resting-state EEG activity across aperiodic and period spectral features. The finding suggest that DS is associated with age-related changes in cortical population activity and disrupted theta-to-alpha maturation. These results extend prior cross-sectional work and support a model in which trisomy 21 alters the coordination of early neural circuit maturation rather than uniformly delaying typical EEG development. Longitudinal studies linking EEG trajectories to language outcomes medical factors, and more direct measures of thalamocortical and inhibitory circuit development may help determine whether EEG spectra profiles can serve as markers of developmental heterogeneity in DS.

## Materials and Methods

### Participants

Participants were drawn from five studies that took place in the Labs of Cognitive Neuroscience at Boston Children’s Hospital. Studies are summarized in Supplement Table 1. All DS participants had confirmed Trisomy 21.

Across all studies, 105 participants with DS and 165 TD participants met inclusion criteria (SI Appendix, Section S1.1) and had at least one EEG session. Individual EEG sessions were excluded if the data did not meet quality metrics (SI Appendix, Section S1.4) resulting in a final sample of 86 participants with DS (EEG n = 120) and 154 TD participants (EEG n = 328). This sample is expanded from our previous study with 29 participants with DS^29^. All participants provided informed consent prior to participation. All studies were reviewed and approved by the Boston Children’s Hospital Institutional Review Board. Written informed consent was obtained from each participant’s legal guardian before participation. In addition, a secondary analysis IRB was approved for combining data across studies (IRB-P00037531).

### Resting State EEG

Across all studies, resting state (non-task) continuous EEG data were collected in a sound attenuated, electrically shielded room with dimmed lighting while participants were seated either independently or on their caregiver’s lap while watching a moving screensaver or preferred silent video of their choosing for 2 to 5 minutes. EEG was recorded using a 128-channel HydroCel Geodesic Sensor Net (Electrical Geodesics Inc., Eugene, OR) connected to a NetAmps 400 amplifier (Electrical Geodesics Inc.) sampled either at 500 Hz or 1000 Hz with online referencing to a single vertex electrode (Cz).

Prior to preprocessing, EEG data from sections or entire recordings with significant sound interference or behavioral non-compliance were not included in subsequent power analysis. (See Supplemental Methods for more details). EEG data was then analyzed using the Batch Automated Processing Platform (BEAPP^48^) with the integrated Harvard Automated Preprocessing Pipeline for EEG (HAPPE v1^49^).

#### EEG Spectral decomposition

Within BEAPP, for each two second segment, power spectral density (PSD) was calculated at each electrode using a multitaper spectral analysis^50^ with seven orthogonal tapers. The PSD was averaged across segments and across regions of interest (SI Appendix, Section S1.3). A modified version of SpecParam v1.0.0^51^; https://github.com/fooof-tools/fooof; Python v3.6.8) was used to parametrize aperiodic and periodic components of the power spectra. See Wilkinson et al 2024^19^ for details on modifications to original algorithm - https://osf.io/u3gp4/).

The final sample showed a mean R² = 0.997 (SD = 0.004, range = 0.962 - 0.999).

The SpecParam model estimates the aperiodic component as L = b - log(k + F^β^), where F is frequency, *b* is the offset, and *β* is the exponent or slope. We calculated aperiodic offset at 2.5 Hz rather than using the algorithm’s default 0 Hz extrapolation as there are high levels of error in the SpecParam estimates below 2.5Hz^19^. The periodic power spectrum was derived by subtracting the estimated aperiodic component from the absolute power spectrum. To facilitate peak detection, we applied smoothing to periodic spectra using a Savitzky-Golay filter (scipy.signal.savgol_filter; window length = 101, polyorder =8). Periodic power was calculated using established frequency ranges: theta (4-6 Hz), low beta (12-20 Hz), and high beta (20-30 Hz), gamma (30-50Hz). Given expected increases in alpha peak frequency with age, the maximum peak between 4 – 12Hz was identified and characterized by peak frequency and amplitude. As multiple participants exhibited double peaks within the 4-12 Hz range, we also identified peaks within a narrow theta (4-6.5 Hz) and alpha (6.5-12 Hz) range as done in prior longitudinal infant studies.

Quality metric comparison revealed some significant differences between groups for most metrics (see Supplemental Table 2). Given group differences, mean retained artifact probability was used as a covariate in subsequent EEG feature models, as it was among the metrics most consistently associated with both diagnostic group and age, with the strongest association for age (q < .0001) relative to ICs rejected (q = .015).

### Statistical Analyses

Group differences in verbal and nonverbal developmental quotient scores and EEG quality characteristics were calculated using independent sample t-tests and differences in demographic measures were computed with chi-square tests.

#### Linear mixed effects models (LMM)

To assess differences in developmental trajectories of EEG features we used a linear mixed effect models (Python *statsmodels* package, *mixedlm*, version 0.14.6) with fixed effects for diagnostic group (DS, TD), age in months centered at 36 months, and their interaction (group × age). Mean artifact probability was z-scored within each EEG feature’s analytic sample and included as a covariate to adjust for EEG quality. Participant was included as a random intercept to account for repeated EEG recordings within children. Models were fit using restricted maximum likelihood.

#### EEG feature ∼ diagnostic group × age centered at 36 months + artifact probability z-score + (1 | participant)

Within-group age-related slopes for TD and DS participants were determined using a post-hoc Wald contrast. To account for multiple comparisons across EEG features, false discovery rate correction was applied using the Benjamini-Hochberg procedure separately for each model term. Model-estimated age trajectories were generated from the fixed effects with artifact probability held at the sample mean. Johnson–Neyman bounds were identified as the ages at which the 95% confidence interval for the DS–TD contrast crossed zero. These analyses were used to describe regions of the observed age range in which the adjusted DS–TD difference was statistically distinguishable from zero.

#### Manifold learning and brain–behavior analyses

Participant matching: One EEG recording per DS participant (earliest visit) was retained and TD timepoints were matched to the DS age distribution by greedy nearest-age selection (target 2:1 TD:DS) in order to better equate the sample distribution and the final sample included in the following analysis was n=238 (TD: 152; DS: 86).

Dimensionality reduction: To reduce dimensionality and minimize collinearity among neighboring frequency bins, periodic spectral features were summarized using band-level principal component analysis (PCA using PCA from scikit-learn) prior to manifold learning. Given the known age-related changes in periodic power and spectral peak characteristics within the theta/alpha and beta frequency ranges in TD children^19^, spectral power features were separated into broad theta-alpha (4-12 Hz) and beta (12-30 Hz) bands, standardized (using StandardScaler from scikit-learn), and analyzed separately using PCA. For each frequency band, we retained the minimum number of principal components that explained at least 90% of the variance. For the broad theta-alpha band, three principal components accounted for 98.0% of the variance. For the beta band, two principal components accounted for 92.3% of the variance. These 5 PCA-derived features were combined with four aperiodic and periodic measures derived from SpecParam analyses, including the aperiodic exponent, aperiodic offset, periodic gamma power, and theta/alpha peak characteristics and z-scored prior to PHATE analyses. Peak characteristics were coded as the presence of a theta peak only, an alpha peak only, or both theta and alpha peaks. This resulted in a final feature vector consisting of nine variables. Given the sample size (n= 238), this yielded a feature-to-sample ratio of approximately 1:26.

PHATE Manifold Learning: To capture potentially nonlinear developmental variation in EEG spectral organization, we applied PHATE (Potential of Heat-diffusion for Affinity-based Transition Embedding^32^; phate package in python), a manifold learning approach that builds upon diffusion maps^52^ and is designed to preserve both local and global structure within high-dimensional data. It has also recently been used to capture nonlinear brain features^53–55^ and its association with cognition^56^. A single two-dimensional PHATE embedding was fit jointly across groups on the nine-feature matrix (k-NN = 5, decay = 40). Parameters for band-level PCA and PHATE are detailed in Supplemental Methods.

Brain-behavior associations were tested using multiple linear regression. Within each group (TD and DS), each PHATE dimension was modeled as a function of verbal developmental quotient (VDQ) and chronological age (PHATE dimension ∼ VDQ + age), yielding four within-group models. Specificity analyses additionally adjusted for nonverbal developmental quotient (NVDQ), with verbal developmental quotient. Specificity analyses additionally adjusted for NVDQ. Statistical significance was assessed using Freedman-Lane permutation tests (5,000 permutations), and Benjamini–Hochberg false discovery rate correction was applied separately for each predictor across the four within-group models (i.e., the four VDQ coefficients as one family and the four age coefficients as another); corrected values are reported as permutation q-values.

## Use of AI

During the preparation of this manuscript, the authors used ChatGPT and Claude to assist in writing Python scripts, as well as to improve clarity and the readability of the Introduction and Discussion while staying in word count limits. All AI-suggested code was reviewed and validated. All AI-suggested text was reviewed by the authors. The authors take full responsibility for the published content.

## Data availability

Consents obtained from human participants prohibit sharing of de-identified individual data without data use agreement in place. Please contact the corresponding author with data requests.

## Acknowledgements

We thank all the children and families who generously participated in this research. We thank all the research staff involved in participant recruitment, data collection, and database administration.

## Author Contributions

**Conceptualization:** CW, NB

**Data curation:** MT, CW

**Formal Analysis:** MT, HC, CW

**Funding Acquisition:** CW, NB

**Investigation:** MT, HC, CW

**Methodology:** HC, CW

**Project Administration:** MT, KP, CW

**Software:** MT, HC, CW

**Supervision:** KP, CW, NB

**Visualization:** MT, HC, CW

**Writing – Original draft:** MT, HC, CW

**Writing – review & editing:** KP, NB

**AI –** CW wrote portions of the manuscript with language editing assistance from ChatGPT (GPT-5.5).

## References

1. Antón-Bolaños, N., Espinosa, A. & López-Bendito, G. Developmental interactions between thalamus and cortex: a true love reciprocal story. Curr. Opin. Neurobiol. 52, 33–41 (2018).

2. Glantz, L. A., Gilmore, J. H., Hamer, R. M., Lieberman, J. A. & Jarskog, L. F. Synaptophysin and postsynaptic density protein 95 in the human prefrontal cortex from mid-gestation into early adulthood. Neuroscience 149, 582–591 (2007).

3. Kim, J.-Y. & Paredes, M. F. Implications of Extended Inhibitory Neuron Development. Int. J. Mol. Sci. 22, 5113 (2021).

4. Lopes da Silva, F. Neural mechanisms underlying brain waves: from neural membranes to networks. Electroencephalogr. Clin. Neurophysiol. 79, 81–93 (1991).

5. Molnár, Z., Luhmann, H. J. & Kanold, P. O. Transient cortical circuits match spontaneous and sensory-driven activity during development. Science 370, eabb2153 (2020).

6. Shen, M. D. et al. Subcortical Brain Development in Autism and Fragile X Syndrome: Evidence for Dynamic, Age- and Disorder-Specific Trajectories in Infancy. Am. J. Psychiatry 179, 562–572 (2022).

7. Pretzsch, C. M. et al. Patterns of Brain Maturation in Autism and Their Molecular Associations. JAMA Psychiatry 81, 1253–1264 (2024).

8. Námešná, A. et al. Synaptic and Circuit Mechanisms Shaping Neurodevelopmental and Psychiatric Outcomes Associated with 16p11.2 Copy Number Variation. Genes 17, 716 (2026).

9. Johnson, M. H., Jones, E. J. H. & Gliga, T. Brain adaptation and alternative developmental trajectories. Dev. Psychopathol. 27, 425–442 (2015).

10. Bull, M. J. Down Syndrome. N. Engl. J. Med. 382, 2344–2352 (2020).

11. Karmiloff-Smith, A. et al. The importance of understanding individual differences in Down syndrome. F1000Research 5, F1000 Faculty Rev-389 (2016).

12. Abbeduto, L., Warren, S. F. & Conners, F. A. Language development in Down syndrome: from the prelinguistic period to the acquisition of literacy. Ment. Retard. Dev. Disabil. Res. Rev. 13, 247–261 (2007).

13. Goodspeed, K. et al. Electroencephalographic (EEG) Biomarkers in Genetic Neurodevelopmental Disorders. J. Child Neurol. 38, 466–477 (2023).

14. Manning, J. R., Jacobs, J., Fried, I. & Kahana, M. J. Broadband shifts in local field potential power spectra are correlated with single-neuron spiking in humans. J. Neurosci. 29, 13613–13620 (2009).

15. Gao, R., Peterson, E. J. & Voytek, B. Inferring synaptic excitation/inhibition balance from field potentials. NeuroImage 158, 70–78 (2017).

16. Voytek, B. et al. Age-related changes in 1/f neural electrophysiological noise. J. Neurosci. 35, 13257–13265 (2015).

17. Buzsáki, G. & Chrobak, J. J. Temporal structure in spatially organized neuronal ensembles: a role for interneuronal networks. Curr. Opin. Neurobiol. 5, 504–510 (1995).

18. Hughes, S. W. & Crunelli, V. Thalamic mechanisms of EEG alpha rhythms and their pathological implications. Neurosci. Rev. J. Bringing Neurobiol. Neurol. Psychiatry 11, 357–372 (2005).

19. Wilkinson, C. L. et al. Developmental trajectories of EEG aperiodic and periodic components in children 2–44 months of age. Nat. Commun. 15, 5788 (2024).

20. Cellier, D., Riddle, J., Petersen, I. & Hwang, K. The development of theta and alpha neural oscillations from ages 3 to 24 years. Dev. Cogn. Neurosci. 50, 100969 (2021).

21. McSweeney, M. et al. Age-related trends in aperiodic EEG activity and alpha oscillations during early- to middle-childhood. NeuroImage 269, 119925 (2023).

22. Stanyard, R. A. et al. Aperiodic and Hurst EEG exponents across early human brain development: A systematic review. Dev. Cogn. Neurosci. 68, 101402 (2024).

23. Sacks, D. D. et al. Longitudinal Trajectories of Aperiodic EEG Activity in Early to Middle Childhood. Child Dev. 96, 1688–1699 (2025).

24. Hill, A. T., Clark, G. M., Bigelow, F. J., Lum, J. A. G. & Enticott, P. G. Periodic and aperiodic neural activity displays age-dependent changes across early-to-middle childhood. Dev. Cogn. Neurosci. 54, 101076 (2022).

25. Marshall, P. J., Bar-Haim, Y. & Fox, N. A. Development of the EEG from 5 months to 4 years of age. Clin. Neurophysiol. 113, 1199–1208 (2002).

26. Tröndle, M., Popov, T., Dziemian, S. & Langer, N. Decomposing the role of alpha oscillations during brain maturation. eLife 11, e77571 (2022).

27. Orekhova, E. V., Stroganova, T. A. & Posikera, I. N. Theta synchronization during sustained anticipatory attention in infants over the second half of the first year of life. Int. J. Psychophysiol. 32, 151172 (1999).

28. Rico-Picó, J. et al. Early development of electrophysiological activity: Contribution of periodic and aperiodic components of the EEG signal. Psychophysiology 60, e14360 (2023).

29. Geiger, M., Hurewitz, S. R., Pawlowski, K., Baumer, N. T. & Wilkinson, C. L. Alterations in aperiodic and periodic EEG activity in young children with Down syndrome. Neurobiol. Dis. 200, 106643 (2024).

30. Babiloni, C. et al. Inter-hemispheric functional coupling of eyes-closed resting EEG rhythms in adolescents with Down syndrome. Clin. Neurophysiol. 120, 1619–1627 (2009).

31. Hamburg, S., Bush, D., Strydom, A. & Startin, C. M. Comparison of resting-state EEG between adults with Down syndrome and typically developing controls. J. Neurodev. Disord. 13, 48 (2021).

32. Moon, K. R. et al. Visualizing structure and transitions in high-dimensional biological data. Nat. Biotechnol. 37, 1482–1492 (2019).

33. Mullen, E. Mullen Scales of Early Learning (AGS Edition). (Circle Pines: American Guidance Service, 1995).

34. Zimmerman, I. L., Steiner, V. G. & Pond, R. E. Preschool Language Scales, Fifth Edition. (Pearson Education, Inc, 2011).

35. Chini, M., Pfeffer, T. & Hanganu-Opatz, I. An increase of inhibition drives the developmental decorrelation of neural activity. eLife 11, e78811 (2022).

36. McKeon, S. D. et al. Aperiodic EEG and 7T MRSI evidence for maturation of E/I balance supporting the development of working memory through adolescence. Dev. Cogn. Neurosci. 66, 101373 (2024).

37. Zorrilla de San Martin, J., Delabar, J.-M., Bacci, A. & Potier, M.-C. GABAergic over-inhibition, a promising hypothesis for cognitive deficits in Down syndrome. Free Radic. Biol. Med. 114, 33–39 (2018).

38. Liu, H. et al. DSCAM gene triplication causes excessive GABAergic synapses in the neocortex in Down syndrome mouse models. PLoS Biol. 21, e3002078 (2023).

39. Souchet, B. et al. Excitation/inhibition balance and learning are modified by Dyrk1a gene dosage. Neurobiol. Dis. 69, 65–75 (2014).

40. Ben-Ari, Y. Excitatory actions of gaba during development: the nature of the nurture. Nat. Rev. Neurosci. 3, 728–739 (2002).

41. Ben-Ari, Y., Gaiarsa, J.-L., Tyzio, R. & Khazipov, R. GABA: a pioneer transmitter that excites immature neurons and generates primitive oscillations. Physiol. Rev. 87, 1215–1284 (2007).

42. Deidda, G. et al. Reversing excitatory GABAAR signaling restores synaptic plasticity and memory in a mouse model of Down syndrome. Nat. Med. 21, 318–326 (2015).

43. Lysenko, L. V. et al. Developmental excitatory-to-inhibitory GABA polarity switch is delayed in Ts65Dn mice, a genetic model of Down syndrome. Neurobiol. Dis. 115, 1–8 (2018).

44. Vuong, C. K. et al. A single-cell multiomic analysis identifies molecular and gene-regulatory mechanisms dysregulated in developing Down syndrome neocortex. Science 392, eaea1259 (2026).

45. Wilkinson, C. Increased aperiodic gamma power in young boys with Fragile X Syndrome is associated with better language ability. 10.21203/rs.3.rs-96363/v2 (2021) doi:10.21203/rs.3.rs-96363/v2.

46. Antoine, M. W., Langberg, T., Schnepel, P. & Feldman, D. E. Increased Excitation-Inhibition Ratio Stabilizes Synapse and Circuit Excitability in Four Autism Mouse Models. Neuron 101, 648–661.e4 (2019).

47. Jones, S. R. When brain rhythms aren’t ‘rhythmic’: implication for their mechanisms and meaning. Curr. Opin. Neurobiol. 40, 72–80 (2016).

48. Levin, A. R., Méndez Leal, A. S., Gabard-Durnam, L. J. & O’Leary, H. M. BEAPP: The Batch Electroencephalography Automated Processing Platform. Front. Neurosci. 12, (2018).

49. Gabard-Durnam, L. J., Mendez Leal, A. S., Wilkinson, C. L. & Levin, A. R. The Harvard Automated Processing Pipeline for Electroencephalography (HAPPE): Standardized Processing Software for Developmental and High-Artifact Data. Front. Neurosci. 10.3389/fnins.2018.00097 (2018) doi:10.3389/fnins.2018.00097.

50. Babadi, B. & Brown, E. N. A review of multitaper spectral analysis. IEEE Trans. Biomed. Eng. 61, 1555–1564 (2014).

51. Donoghue, T. et al. Parameterizing neural power spectra into periodic and aperiodic components. Nat. Neurosci. 23, 1655–1665 (2020).

52. Coifman, R. R. et al. Geometric diffusions as a tool for harmonic analysis and structure definition of data: diffusion maps. Proc. Natl. Acad. Sci. U. S. A. 102, 7426–7431 (2005).

53. De, A. & Chaudhuri, R. Common population codes produce extremely nonlinear neural manifolds. Proc. Natl. Acad. Sci. U. S. A. 120, e2305853120 (2023).

54. Gao, S., Mishne, G. & Scheinost, D. Nonlinear manifold learning in functional magnetic resonance imaging uncovers a low-dimensional space of brain dynamics. Hum. Brain Mapp. 42, 4510–4524 (2021).

55. Busch, E. L., Fincke, E. C., Lajoie, G., Krishnaswamy, S. & Turk-Browne, N. B. Human learning of noninvasive brain-computer interfaces via manifold geometry. Nat. Neurosci. 10.1038/s41593-026-02311-2 (2026) doi:10.1038/s41593-026-02311-2.

56. Busch, E. L. et al. Multi-view manifold learning of human brain-state trajectories. Nat. Comput. Sci. 3, 240–253 (2023).

