## Supplemental Appendix for "Altered early cortical EEG maturation and its relationship to language development in Down syndrome"

**(1) SUPPLEMENTAL METHODS**

**1.1 Participant Inclusion:** This analysis includes infants and children aged 12-82 months with either Down syndrome (DS), or without DS and no known developmental delays (termed "Typically Developing (TD) Group"). Participants were drawn from the following five studies summarized in Supplemental Table 1: (1) Infant Screening Project (IRB-P00018377; TD and DS), (2) The Joint Attention, Symbolic Play, Engagement and Regulation (JASPER) Project in Down Syndrome (IRB-P00025806; DS only), (3) Fragile X Neural Markers Study (IRB-P00025493; TD only), (4) Brain Indicators of Developmental Growth (IRB-P00034676; DS & TD), and (5) EEG Collection for Pilot Projects (IRB-P00048681; DS only). Analysis for this paper is specific to those older than 12 months of age and either with DS diagnosis or TD. Across all five studies and both groups, participants were excluded from analysis if they had a known neurological disorder (except a diagnosis of DS), including but not limited to intraventricular hemorrhage or unstable seizures. Additionally, participants were excluded if the child had a history of birth trauma, current use of anticonvulsant medications, or prenatal opioid exposure. TD participants were also excluded if they had a diagnosis of autism spectrum disorder, known genetic disorder, or cognitive delay. All DS participants had confirmed Trisomy 21.

Given the high prevalence of co-occurring health and cognitive conditions in DS populations, we did not exclude for minimal-moderate hearing loss or eustachian tube dysfunction (DS = 39, TD = 7), visual impairment (e.g., wears glasses, has astigmatism) (DS = 39, TD = 8) or other frequently associated conditions including sleep disorders, cardiovascular diseases, and gastrointestinal issues. All data used in the present study were approved by the institutional review.

**Supplemental Table 1**

| | Study 1 | Study 2 | Study 3 | Study 4 | Study 5 |
|---|---|---|---|---|---|
| Analysis Group | DS (n = 23)<br>TD (n = 94) | DS (n = 11) | TD (n = 14) | DS (n = 32)<br>TD (n = 51) | DS (n = 29) |
| Age Range | 12 – 39 months | 29 – 48 months | 32 – 80 months | 24 – 82 months | 12 – 75 months |
| Min. Gestational Age | ≥34 weeks | ≥33 weeks | >35 weeks | >35 weeks | >36 weeks |
| English Language Requirement | > 50% heard of waking hours | > 50% heard of waking hours | > 50% heard of waking hours | > 75% heard of waking hours | None |
| Data collection | Longitudinal (TD)<br>Cross-sectional (DS) | Cross-sectional | Cross-sectional | Longitudinal | Cross-sectional and Longitudinal |
| Developmental Assessments | Mullen Scales of Early Learning | Mullen Scales of Early Learning | Mullen Scales of Early Learning | Preschool Language Scale-5 | None |

**1.2 EEG event tagging**

Raw EEG data from studies 3-5 were event tagged for significant sound interference (video sound on, adult or participant vocalizations/talking) or behavioral non-compliance based on reviewing of the time-locked video recording of the EEG session. Segments of EEG data that were tagged for sound or behavioral disruptions were excluded from subsequent power analyses. The EEG data of studies 1 and 2 were not tagged for such events, however, for all studies if experimenters rated behavioral compliance on a general scale of either 1 to 5 or 1 to 10, EEG sessions that had a rating of 1 were excluded from the analysis. To further assess potential differences in data quality between event tagged or raw EEG data, HAPPE quality metrics were compared across studies 1-2 and studies 3-5. Linear mixed-effects models controlling for enrollment group and age, with FDR correction applied across quality metrics, revealed that event-tagged and raw data differed only in their number of retained segments ($\beta = -11.34$, 95% CI [−18.62, −4.06], $p = .002$, $q = .011$), with event-tagged data retaining fewer segments on average than raw data. This decrease in number of kept segments is expected as event tagging and subsequent removal of sound- and behavior-tagged segments necessarily reduces the total amount of retained data relative to sessions without event-level tagging. Groups did not differ significantly on any other quality metrics, including the proportion of good channels, proportion of independent components (ICs) rejected, mean artifact probability of retained ICs, or percentage of EEG variance retained (all $q > .05$) (See Supplementary Table 3)

#### 1.3 EEG preprocessing

EEG data from Netstation files (Electrical Geodesics, Inc.) were exported to MATLAB (version R2021a) for pre-processing and power spectrum calculations using the Batch Automated Processing Platform (BEAPP[1]) with the integrated Harvard Automated Preprocessing Pipeline for EEG (HAPPE v1[2]). Each EEG recording underwent bandpass filtering with high-pass and low-pass cutoffs at 1 Hz and 100 Hz, respectively. Data originally sampled at 1000 Hz or 500 Hz were down-sampled to 250 Hz before processing through the HAPPE module. The HAPPE v1 pipeline performed 60 Hz line noise removal, bad channel identification and rejection, and artifact removal using combined wavelet-enhanced independent component analysis (ICA) and the Multiple Artifact Rejection Algorithm (MARA[3,4]). Due to the brief duration of EEG recordings, a pre-selected subset of channels providing even coverage across all brain regions of interest was used for ICA/MARA based artifact removal: (Standard 10–20 electrodes:

Supplemental Figure 1

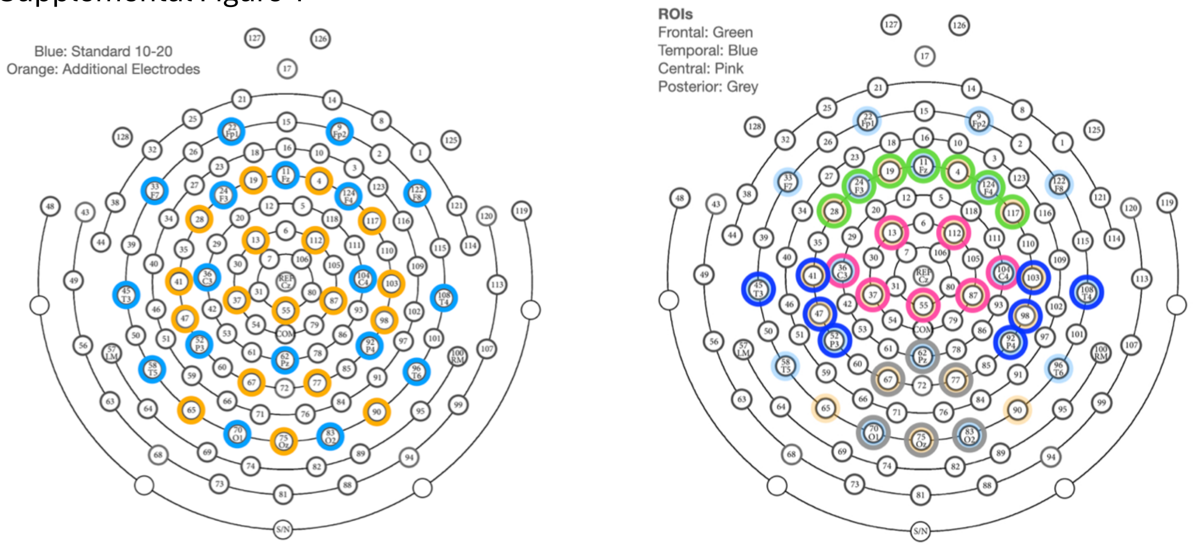


9, 11, 22, 24, 33, 36, 45, 52, 58, 62, 70, 83, 92, 96, 104, 108, 122, 124; Additional electrodes: 4, 13, 19, 28, 37, 41, 47, 55, 65, 67, 75, 77, 87, 90, 98, 103, 112, 117; Supplementary Figure 1). Following artifact removal, channels that were rejected during bad channel detection were interpolated. Data were then re-referenced to the average reference and detrended relative to the signal mean. Finally, data was segmented into 2-second epochs and any segments containing residual artifacts were identified and rejected using HAPPE's amplitude and joint probability criteria to ensure data quality for subsequent analyses.

SpecParam was run in the fixed mode without a spectral knee across the 2 to 55 Hz frequency range. Parameters included: *peak_width_limits* were set to [0.5, 18.0], *max_n_peak* = 7 peaks, and *peak_threshold* = 2.

**1.4 EEG exclusions criteria and quality metrics:** EEG data were excluded from final analysis if they met any of the following criteria: fewer than 20 good segments, percent good channels < 80%, percent independent components (ICs) rejected as artifact > 80%, mean retained artifact probability > 0.3, or percent of EEG signal variance retained after artifact removal < 25% (DS: n = 6, TD: n = 8). Finally, recordings were excluded if the mean SpecParam model fit quality, averaged across all electrodes and regions of interest, fell below an $R^2$ of 0.97 (DS: n = 2, TD  n = 3). Given repeated measures and differences in age distribution between groups, group differences in EEG quality were assessed using linear mixed models (Supplemental Table 2):

*EEG quality metric ~ diagnostic group + age centered at 36 months + (1 | participant)*

As no group × age interaction was included, the group coefficient reflects the adjusted DS –TD difference in each quality metric. Benjamini-Hochberg false discovery rate correction was applied for each model term across all quality metrics.

**Supplemental Table 2** EEG Quality Metrics

| | **TD mean at 36 mo [95% CI]** | **DS mean at 36 mo [95% CI]** | **DS vs TD** |
|---|---|---|---|
| Number of kept segments | 103.43 [99.61, 107.25] | 93.03 [87.33, 98.73] | −10.40 [−17.21, −3.59], *p* = .003, q = .005 |
| Proportion Good channels | 0.94 [0.93, 0.94] | 0.93 [0.93, 0.94] | −0.00 [−0.01, 0.01], *p* = .355, q = .355 |
| Proportion ICs rejected | 0.34 [0.33, 0.36] | 0.42 [0.40, 0.44] | 0.08 [0.05, 0.10], *p* < .0001, q < .0001 |
| Mean retained artifact probability | 0.108 [0.102, 0.114] | 0.140 [0.132, 0.149] | 0.03 [0.02, 0.04], *p* < .0001, q < .0001 |
| % EEG signal variance retained | 69.00 [66.85, 71.15] | 64.38 [61.27, 67.50] | −4.61 [−8.39, −0.83], *p* = .017, q = .021 |

Age (centered at 36 months) was as a covariate though it is not shown. Means are estimated by model at 36 months.

**Supplementary Table 3** EEG Quality Metrics Between Event-Tagged and Raw Data Groupings

| | **Event-Tagged (n= 155) vs Raw (n= 293)** | **Enrollment Group Effect** | **Age Effect** |
|---|---|---|---|
| Number of kept segments | −11.34 [−18.62, −4.06], p = .002, q = .011 | −8.05 [−14.90, −1.21], *p* = .021, q = .035 | 2.13 [−0.23, 4.49], *p* = .077, q = .129 |
| Good channels (%) | 0.00 [−0.01, 0.01], *p* = .442, q = .442 | −0.01 [−0.02, 0.00], *p* = .283, q = .283 | −0.00 [−0.00, 0.00], *p* = .589, q = .589 |

| | | | |
|---|---|---|---|
| ICs rejected (%) | 0.02 [−0.01, 0.05], *p* = .116, q = .201 | 0.07 [0.04, 0.10], *p* < .0001, q < .0001 | −0.01 [−0.02, −0.01], *p* = .001, q = .004 |
| Mean retained artifact probability | −0.01 [−0.02, 0.00], *p* = .120, q = .201 | 0.03 [0.02, 0.05], *p* < .0001, q < .0001 | −0.01 [−0.01, −0.00], *p* = .002, q = .004 |
| EEG signal variance retained (%) | −2.51 [−6.60, 1.58], *p* = .229, q = .286 | −4.07 [−7.96, −0.19], *p* = .040, q = .050 | 0.72 [−0.52, 1.96], *p* = .256, q = .320 |

**Supplemental Figure 2:**
(A and B) Mean aperiodic fit (blue), Absolute spectrum (red), and FOOOF estimated spectrum (green) for children with DS (A) or typically developmental children (B). (C) Mean absolute error of FOOOF/SpecParam fit For children with DS (blue) or TD (orange).

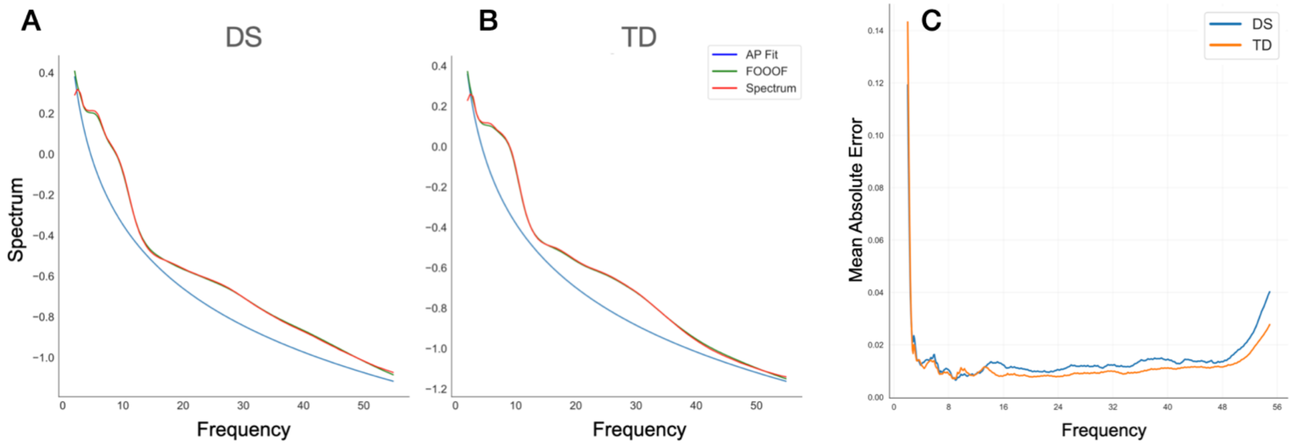


### 1.5 Developmental Assessments

Across studies, a nonverbal developmental quotient (NVDQ) was determined using the Mullen Scales of Early Learning (MSEL[5]). The MSEL is a standardized developmental measure validated for children between the ages of 0 and 69 months. NVDQ was calculated using average the age equivalents for the visual receptions and fine motor subscales of the MSEL, divided by chronological age in months and multiplied by 100.

Verbal developmental quotient (VDQ) was determined using either age-equivalents on expressive and receptive language subscales of the MSEL (Studies 1-3) or on the expressive communication and auditory comprehension subscales of the Preschool Language Scale-5 (PLS-5[6]) (Studies 4 & 5). The PLS-5 is standardized language assessment validated for children between the ages of 0 and 95 months. Age, NVDQ, and VDQ are summarized across all available data, thus including multiple observations per participant where applicable, rather than being limited to a single observation per participant (Supplemental Table 4).

**Supplementary Table 4** Verbal and Non-verbal Developmental Quotient Grouping

| | DS | | | TD | | |
|---|---|---|---|---|---|---|
| | ***n*** | **M (SD) Range** | ***Range*** | ***n*** | **M (SD)** | ***Range*** |
| Age at visit | 86 | 40.44 (15.10) | 13.90 - 81.64 | 320 | 29.64 (16.42) | 12.02 - 78.52 |
| VDQ | 85 | 52.23 (13.67) | 23.96 - 89.47 | 309 | 109.72 (14.94) | 63.59 - 145.83 |
| NVDQ | 85 | 51.95 (13.96) | 8.89 - 81.25 | 320 | 112.66 (18.33) | 61.54 - 154.17 |

#### 1.6 Manifold learning and brain–behavior analyses

Dimensionality reduction: To reduce dimensionality and minimize collinearity among neighboring frequency bins, periodic spectral features were summarized using band-level principal component analysis (PCA; using PCA from scikit-learn) prior to manifold learning. Given known age-related changes in periodic power and spectral peak characteristics within the theta/alpha and beta frequency ranges in TD children[7], spectral power features were separated into broad theta-alpha (4-12 Hz) and beta (12-30 Hz) bands, standardized (using StandardScaler from scikit-learn), and analyzed separately using PCA. For each frequency band, we retained the minimum number of principal components that explained at least 90% of the variance. For the broad theta-alpha band, three principal components accounted for 98.0% of the variance. For the beta band, two principal components accounted for 92.3% of the variance. (Supplemental Figure 3).

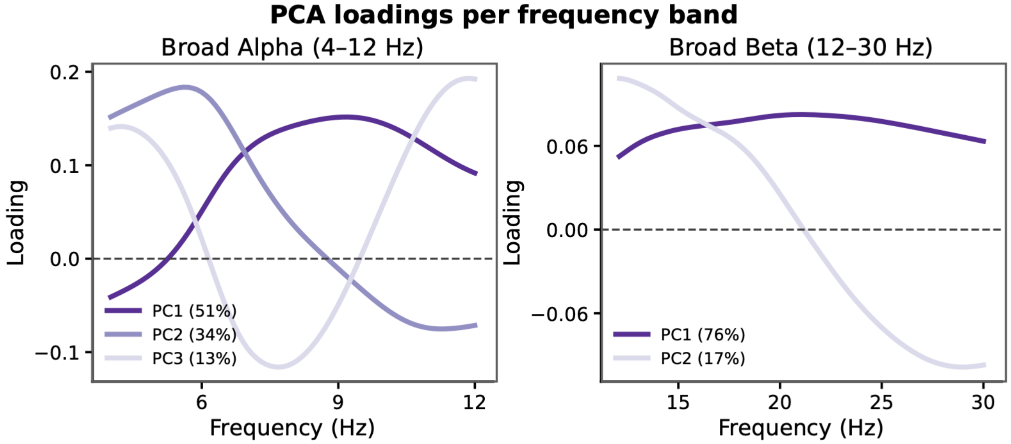


**Supplemental Figure 3.** Principal component analysis (PCA) of the broad theta-alpha band (4–12 Hz), showing the loading profiles of the three retained principal components (left) and the broad beta band (12–30 Hz), showing the loading profiles of the two retained principal components (right).

PHATE manifold embedding: To capture potentially nonlinear developmental variation in EEG spectral organization, we applied PHATE (phate package in python), a manifold learning approach that preserves both local and global structure within high-dimensional data. PHATE builds upon diffusion maps[8] and is designed to preserve both local and global structure within high-dimensional data. It has also recently been used to capture nonlinear brain features[9–11] and its association with cognition[12]. A k-nearest neighbor graph was constructed using k = 5, and the diffusion operator decay parameter was set to 40. The embedding was computed in a two-dimensional space and used for visualization and subsequent brain-behavior analyses.

### (2) SUPPLEMENTAL RESULTS

**2.1 Income sensitivity analysis:** To assess whether differences in group demographics impacted our analyses of group differences in EEG-feature age associations, we repeated linear mixed-effect models now adding household income as an additional categorical covariate. Household income was modeled using three categories: less than $40,000, $40,000–$69,999, and $70,000 or greater. Participants with missing income data or who preferred not to disclose income were excluded from this sensitivity analysis. Models included fixed effects for diagnostic group, age centered at 36 months, the group-by-age interaction, household income, and artifact probability z-score. Participant was included as a random intercept to account for repeated EEG recordings within children. Models were fit using restricted maximum likelihood.

EEG feature ~ diagnostic group × age centered at 36 months + household income + artifact probability z-score + (1 | participant)

**Supplemental Table 5** EEG-feature Age Associations with Income Covariate

| Income Sensitivity Model | Group effect at 36m β [95% CI], q | Group x Age Interaction β [95% CI], q | TD slope β/year [95% CI], p | DS slope β/year [95% CI], p |
|---|---|---|---|---|
| EEG Feature | | | | |
| Aperiodic Offset | 0.03 [−0.01, 0.07] q = .188 | 0.0 [−0.02, 0.03] q = .895 | −0.00 [−0.01, 0.01] p = .587 | −0.00 [−0.02, 0.02] p = .981 |
| Aperiodic Exponent | 0.01 [−0.02, 0.03] q = .650 | −0.02 [−0.04, −0.00] q = .056 | −0.01 [−0.02, 0.00] p = .085 | −0.03 [−0.04, −0.01] **p < .001** |
| 4-12Hz Peak Frequency | −0.74 [−1.05, −0.43] **q < .0001** | −0.29 [−0.49, −0.10] **q = .012** | 0.44 [0.34, 0.54] **p < .0001** | 0.15 [−0.02, 0.31] p = .083 |
| 4-12Hz Peak Amplitude | −0.01 [−0.03, 0.02] q = .532 | −0.02 [−0.04, −0.01] **q = .004** | 0.01 [0.01, 0.02] **p < .001** | −0.01 [−0.02, −0.00] **p = .042** |
| Periodic Theta Power | 0.14 [0.10, 0.17], **q < .0001** | −0.00 [−0.02, 0.02] q = .895 | −0.03 [−0.04, −0.02] **p < .0001** | −0.04 [−0.05, −0.02] **p < .0001** |
| Periodic Low Beta Power | −0.25 [−0.41, −0.09] **q = .005** | −0.07 [−0.16, 0.02], q = .271 | 0.14 [0.10, 0.19] **p < .0001** | 0.07 [−0.00, 0.15] p = .058, |
| Periodic High Beta Power | −0.15 [−0.39, 0.09] q = .284 | 0.07 [−0.06, 0.20], q = .435 | −0.15 [−0.22, −0.09] **p < .0001** | −0.08 [−0.19, 0.03], p = .151 |
| Periodic Gamma Power | 0.60 [0.30, 0.91] **q < .001** | −0.02 [−0.20, 0.15] q = .895 | −0.39 [−0.48, −0.31] **p < .0001** | −0.42 [−0.57, −0.27] **p < .0001** |

**2.2 EEG Power Spectra across different ROIs**

Aperiodic and periodic spectra across frontal, temporal, central, and posterior ROIs (Supplemental Figure 1) are shown in Supplemental Figure 4.

Linear mixed-effects models examined age-related trajectories of EEG features measured across different electrode-based regions of interest (Supplemental Table 6). EEG quality metric, mean retained artifact probability, was included as a covariate. Diagnostic group, age in months centered at 36 months, and their interaction were included as fixed effects. Participant was included as a random intercept to account for repeated EEG recordings within children. Models were fit using restricted maximum likelihood. Effects were generally similar across regions of interests for most EEG features.

### Supplemental Figure 4

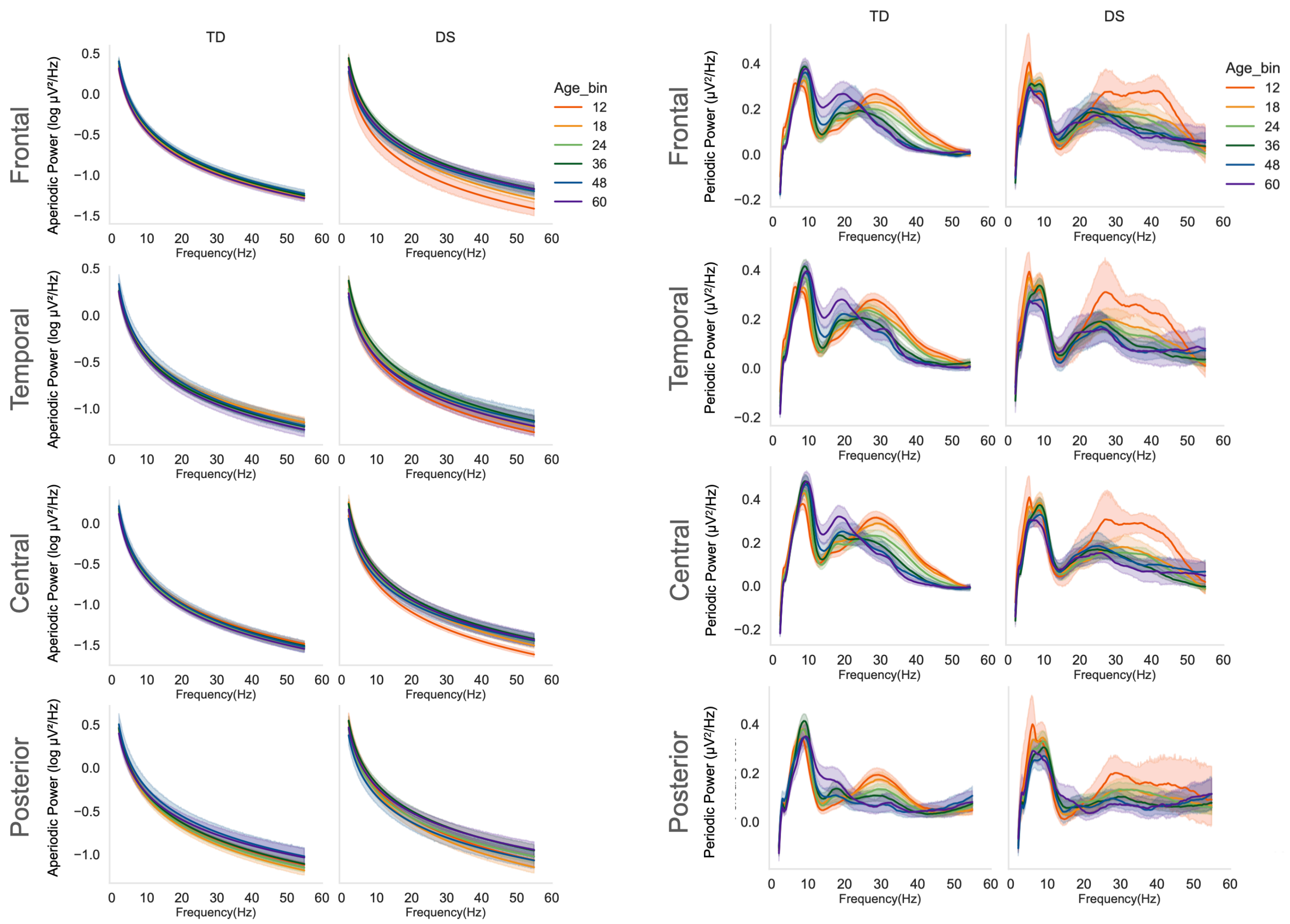


### **Supplemental Table 6** Linear Mixed-Effect Models of Additional ROIs

| | Group effect at 36m β [95% CI], q | Group x Age Interaction β [95% CI], q | TD slope β/year [95% CI], p | DS slope β/year [95% CI], p |
|---|---|---|---|---|
| Aperiodic Offset | | | | |
| Frontal | 0.02 [-0.01, 0.06], q = .294 | -0.01 [-0.03, 0.02], q = .757 | 0.01 [-0.00, 0.02], p = .193 | 0.00 [-0.02, 0.02], p = .776 |
| Temporal | 0.05 [0.01, 0.10], **q = .025** | -0.00 [-0.03, 0.02], q = .747 | 0.00 [-0.01, 0.01], p = .948 | -0.00 [-0.03, 0.02], p = .735 |
| Central | 0.04 [-0.01, 0.09], q = .112 | 0.01 [-0.02, 0.03], q = .725 | -0.01 [-0.02, 0.01], p = .338 | -0.00 [-0.03, 0.02], p = .881 |
| Posterior | 0.03 [-0.01, 0.08], q = .199 | -0.00 [-0.03, 0.03], q = .867 | -0.01 [-0.02, 0.01], p = .236 | -0.01 [-0.04, 0.01], p = .369 |
| Aperiodic Exponent | | | | |
| Frontal | -0.01 [-0.03, 0.02], q = .641 | -0.02 [-0.04, -0.01], **q = .012** | 0.00 [-0.01, 0.01], p = .728 | -0.02 [-0.04, -0.01], **p = .001** |
| Temporal | 0.03 [0.00, 0.06], **q = .025** | -0.02 [-0.04, -0.00], **q = .043** | 0.01 [0.00, 0.02], **p = .009** | -0.01 [-0.03, 0.01], p = .204 |
| Central | -0.00 [-0.02, 0.02], q = .948 | -0.02 [-0.03, -0.00], q = .080 | -0.00 [-0.01, 0.01], p = .530 | -0.02 [-0.03, -0.01], **p = .005** |
| Posterior | 0.00 [-0.03, 0.04], q = .837 | 0.00 [-0.02, 0.03], q = .867 | -0.03 [-0.04, -0.02], **p < .0001** | -0.03 [-0.05, -0.01], **p = .007** |

| | | | | |
|---|---|---|---|---|
| 4-12Hz Peak Frequency | | | | |
| Frontal | -0.87 [-1.17, -0.57], **q < .0001** | -0.21 [-0.40, -0.01], q = .102 | 0.43 [0.33, 0.53], **p < .0001** | 0.22 [0.05, 0.39], **p = .009** |
| Temporal | -0.59 [-0.90, -0.28], **q < .001** | -0.29 [-0.49, -0.10], **q = .015** | 0.52 [0.42, 0.62], **p < .0001** | 0.22 [0.06, 0.39], **p = .009** |
| Central | -0.49 [-0.75, -0.22], **q < .001** | -0.27 [-0.43, -0.10], **q = .008** | 0.35 [0.26, 0.44], **p < .0001** | 0.09 [-0.06, 0.23], p = .245 |
| Posterior | -0.62 [-0.92, -0.32], **q < .001** | -0.29 [-0.48, -0.10], **q = .018** | 0.37 [0.28, 0.47], **p < .0001** | 0.08 [-0.08, 0.24], p = .317 |
| 4-12Hz Peak Amplitude | | | | |
| Frontal | -0.01 [-0.03, 0.01], q = .412 | -0.03 [-0.04, -0.01], **q = .003** | 0.01 [0.01, 0.02], **p < .001** | -0.01 [-0.03, -0.00], **p = .031** |
| Temporal | -0.02 [-0.04, 0.01], q = .225 | -0.03 [-0.05, -0.02], **q < .0001** | 0.02 [0.01, 0.03], **p < .0001** | -0.02 [-0.03, -0.00], **p = .012** |
| Central | -0.05 [-0.07, -0.02], **q < .001** | -0.03 [-0.05, -0.01], **q = .003** | 0.02 [0.01, 0.03], **p < .0001** | -0.01 [-0.02, 0.00], p = .124 |
| Posterior | -0.03 [-0.05, 0.00], q = .125 | -0.02 [-0.04, -0.00], q = .109 | 0.00 [-0.00, 0.01], p = .339 | -0.01 [-0.03, -0.00], **p = .044** |
| Periodic Theta Power | | | | |
| Frontal | 0.15 [0.12, 0.18], **q < .0001** | -0.00 [-0.02, 0.02], q = .757 | -0.04 [-0.05, -0.03], **p < .0001** | -0.04 [-0.06, -0.02], **p < .0001** |
| Temporal | 0.10 [0.07, 0.14], **q < .0001** | -0.01 [-0.03, 0.01], q = .678 | -0.04 [-0.05, -0.03], **p < .0001** | -0.05 [-0.06, -0.03], **p < .0001** |
| Central | 0.11 [0.08, 0.14], **q < .0001** | -0.01 [-0.03, 0.01], q = .518 | -0.02 [-0.03, -0.01], **p < .0001** | -0.03 [-0.05, -0.02], **p < .001** |
| Posterior | 0.11 [0.08, 0.14], **q < .0001** | -0.01 [-0.03, 0.01], q = .408 | -0.02 [-0.03, -0.01], **p < .001** | -0.03 [-0.05, -0.02], **p < .001** |
| Periodic Low Beta Power | | | | |
| Frontal | -0.32 [-0.49, -0.14], **q < .001** | -0.06 [-0.16, 0.04], q = .417 | 0.17 [0.12, 0.22], **p < .0001** | 0.11 [0.02, 0.20], **p = .017** |
| Temporal | -0.37 [-0.54, -0.19], **q < .001** | -0.10 [-0.21, 0.01], q = .140 | 0.16 [0.11, 0.22], **p < .0001** | 0.06 [-0.03, 0.16], p = .169 |
| Central | -0.57 [-0.75, -0.39], **q < .0001** | -0.06 [-0.17, 0.05], q = .350 | 0.14 [0.08, 0.19], **p < .0001** | 0.08 [-0.01, 0.17], p = .094 |
| Posterior | -0.19 [-0.36, -0.01], q = .067 | -0.06 [-0.16, 0.05], q = .449 | 0.12 [0.07, 0.17], **p < .0001** | 0.06 [-0.03, 0.15], p = .185 |
| Periodic High Beta Power | | | | |
| Frontal | -0.23 [-0.49, 0.03], **q = .141** | 0.12 [-0.03, 0.27], q = .264 | -0.09 [-0.16, -0.01], **p = .024** | 0.03 [-0.10, 0.16], p = .672 |
| Temporal | -0.31 [-0.57, -0.04], **q = .025** | 0.08 [-0.08, 0.24], q = .521 | -0.15 [-0.23, -0.07], **p < .001** | -0.07 [-0.20, 0.07], p = .325 |
| Central | -0.57 [-0.85, -0.30], **q < .0001** | 0.17 [0.01, 0.33], q = .080 | -0.20 [-0.28, -0.12], **p < .0001** | -0.03 [-0.17, 0.11], p = .659 |
| Posterior | -0.12 [-0.36, 0.12], q = .375 | 0.06 [-0.08, 0.21], q = .528 | -0.20 [-0.28, -0.13], **p < .0001** | -0.14 [-0.26, -0.02], **p = .024** |
| Periodic Gamma Power | | | | |
| Frontal | 0.98 [0.64, 1.31], **q < .0001** | 0.09 [-0.11, 0.29], q = .527 | -0.58 [-0.68, -0.47], **p < .0001** | -0.49 [-0.66, -0.32], **p < .0001** |
| Temporal | 0.48 [0.16, 0.80], **q = .006** | -0.04 [-0.24, 0.15], q = .746 | -0.43 [-0.53, -0.33], **p < .0001** | -0.47 [-0.64, -0.31], **p < .0001** |
| Central | 0.40 [0.05, 0.75], **q = .035** | 0.21 [-0.01, 0.44], q = .094 | -0.65 [-0.76, -0.53], **p < .0001** | -0.43 [-0.62, -0.24], **p < .0001** |
| Posterior | 0.38 [0.06, 0.69], **q = .048** | -0.14 [-0.34, 0.06], q = .408 | -0.18 [-0.28, -0.08], **p < .001** | -0.32 [-0.50, -0.15], **p < .001** |

### 2.3 Robustness of DS: PHATE 1 and VDQ association

Robustness of the DS verbal association was assessed by recomputing the two-dimensional embedding across PHATE hyperparameters (k-nearest neighbors = 4, 5, and 8; decay = 20, 40, and 60). For each embedding, we tested the partial association between each embedding dimension and VDQ within DS, controlling for age, using permutation analysis. Across all PHATE parameterizations, dimension 1 showed a consistent positive association with verbal developmental quotient (VDQ) in the DS group after adjusting for age (standardized β = 0.446-0.451, permutation p = .008-.011), whereas embedding dimension 2 was not associated with VDQ (standardized β = -0.084 to -0.042, all p > .65).

**Supplemental Figure 5. Permutation analyses for PHATE-behavior associations.** Permutation analyses for PHATE-behavior associations. Null distributions of regression coefficients (β) obtained from 5,000 permutation tests for the primary and secondary analyses. Red vertical lines indicate the observed regression coefficient, whereas gray histograms represent the null distribution expected under random permutation of the outcome. Dashed red lines denote the two-sided significance threshold. The first row shows within-group associations between PHATE 1, VDQ and age. The second row shows the corresponding analyses with PHATE 2. The third row presents permutation tests of the interaction between PHATE dimensions and diagnostic group (DS versus TD). The final row evaluates whether the association between PHATE 1 and VDQ is specific to VDQ accounting for NVDQ in the DS group. Empirical permutation p-values are reported above each panel.

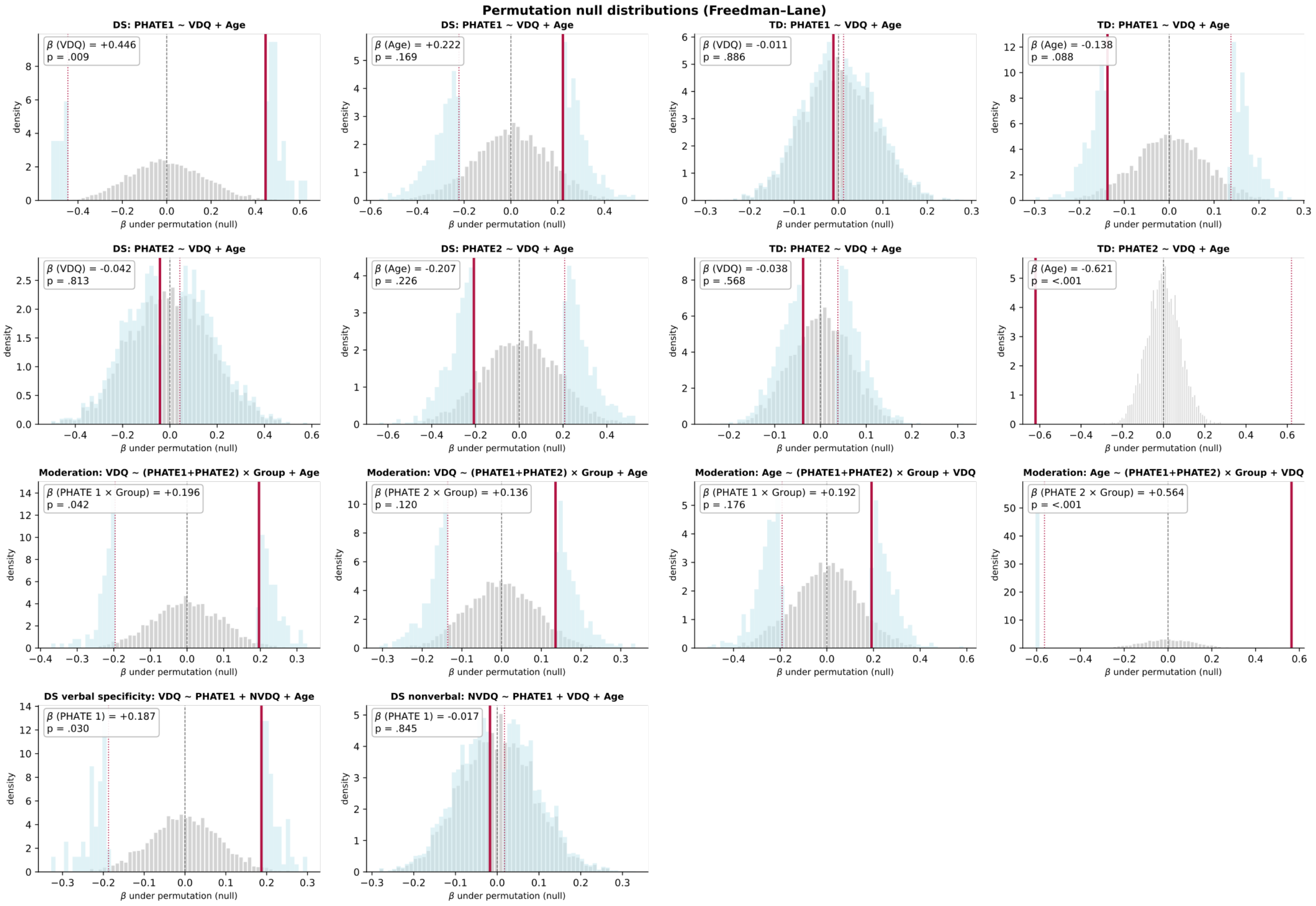

### REFERENCES


1. Levin, A. R., Méndez Leal, A. S., Gabard-Durnam, L. J. & O'Leary, H. M. BEAPP: The Batch Electroencephalography Automated Processing Platform. *Front. Neurosci.* **12**, (2018).
2. Gabard-Durnam, L. J., Mendez Leal, A. S., Wilkinson, C. L. & Levin, A. R. The Harvard Automated Processing Pipeline for Electroencephalography (HAPPE): standardized processing software for developmental and high-artifact data. *Front. Neurosci.* **12**, 97 (2018).
3. Winkler, I., Haufe, S. & Tangermann, M. Automatic Classification of Artifactual ICA-Components for Artifact Removal in EEG Signals. *Behav. Brain Funct.* **7**, 30 (2011).
4. Winkler, I. *et al.* Robust artifactual independent component classification for BCI practitioners. *J. Neural Eng.* **11**, 035013 (2014).
5. Mullen, E. *Mullen Scales of Early Learning (AGS Edition)*. (Circle Pines: American Guidance Service, 1995).
6. Zimmerman, I. L., Steiner, V. G. & Pond, R. E. *Preschool Language Scales, Fifth Edition*. (Pearson Education, Inc, 2011).
7. Wilkinson, C. L. *et al.* Developmental trajectories of EEG aperiodic and periodic components in children 2–44 months of age. *Nat. Commun.* **15**, 5788 (2024).
8. Coifman, R. R. *et al.* Geometric diffusions as a tool for harmonic analysis and structure definition of data: diffusion maps. *Proc. Natl. Acad. Sci. U. S. A.* **102**, 7426–7431 (2005).
9. De, A. & Chaudhuri, R. Common population codes produce extremely nonlinear neural manifolds. *Proc. Natl. Acad. Sci. U. S. A.* **120**, e2305853120 (2023).
10. Gao, S., Mishne, G. & Scheinost, D. Nonlinear manifold learning in functional magnetic resonance imaging uncovers a low-dimensional space of brain dynamics. *Hum. Brain Mapp.* **42**, 4510–4524 (2021).
11. Busch, E. L., Fincke, E. C., Lajoie, G., Krishnaswamy, S. & Turk-Browne, N. B. Human learning of noninvasive brain-computer interfaces via manifold geometry. *Nat. Neurosci.* https://doi.org/10.1038/s41593-026-02311-2 (2026) doi:10.1038/s41593-026-02311-2.
12. Busch, E. L. *et al.* Multi-view manifold learning of human brain-state trajectories. *Nat. Comput. Sci.* **3**, 240–253 (2023).
